# Abrupt increase in the UK coronavirus death-case ratio in December 2020

**DOI:** 10.1101/2021.01.21.21250264

**Authors:** David J Wallace, Graeme J Ackland

## Abstract

1

**Objective:** to determine the statistical relationship between reported deaths and infections in the UK coronavirus outbreak

**Design:** Publicly available UK government data is used to determine a relationship between reported cases and deaths, taking into account various UK regions, age profiles and prevalence of the variant of concern (VOC) B.1.1.7.

**Main Outcome Measures:** Establishing a simple statistical relationship between detected cases and subsequent mortality.

**Results:** Throughout October and November 2020, deaths in England are well described as 1/55^th^ of detected cases from 12 days previously. After that, the relationship no longer holds and deaths are significantly higher. This is especially true in regions affected by the VOC B.1.1.7

**Conclusions:** In early December, some new factor emerged to increase the case-fatality rate in the UK.

**Summary Box:** *What is already known on this topic:* The infection-mortality ratio enables one to predict future deaths based on current infections. Incomplete monitoring of infection may be sufficient to predict future trends.

*What the study adds:* For the specific case of the second wave of coronavirus infection in the UK, we show a clear mathematical relationship between detected infections (positive tests) and subsequent deaths. This relationship begins to fail in December, with unexpectedly high death rates. This may be correlated in time and region with the emergence of the Variant of Concern B 1.1.7.

## 2 Introduction

The ongoing coronavirus crisis needs no introduction. To make plans even a few weeks ahead, it is still important to have some model for how healthcare demands will develop. In November 2020, David Spiegelhalter suggested a simple rule of thumb that on average roughly 1 in 50 positive tests for Covid-19 results in death in 3-4 weeks. This idea projected that Covid-related deaths in the UK would peak at around 450 per day by the end of November, significantly less than many epidemiological projections, and rather accurate, as it transpired. Spiegelhalter’s “model” involves two parameters – the ratio of positive tests to deaths, and a time delay. It should be noted that an exponentially increasing epidemic can be described by a single parameter; the time delay and case-fatality ratios are equivalent. Such independent fitting requires a more distinguishing feature; by early November, the number of positive test peaked, with a subsequent peak in deaths in mid November.

There are several choices of data for deaths and for positive test results. Covid-related deaths are reported in two ways. One is in the headline numbers published daily by the UK and devolved governments which have come in the previous 24 hours; the other is derived from death certificates, giving the number of deaths by date of death. Reporting of the former is highly variable over the week, because of under-reporting during the weekend. This can be smoothed out by taking 7-day rolling averages. For the data in the charts below, these averages are ‘centred’ i.e. include the three days on either side of the date shown. In addition to the weekend effect, UK data was highly skewed by underreporting over Christmas and New Year. Figure 2 shows that published deaths are clearly underreported over Christmas, and catch up again in New Year. To avoid this artefact, data for “date of death” should be used; the last five days of reporting are not used in the charts below, because of incomplete reporting.

**Figure 1.**
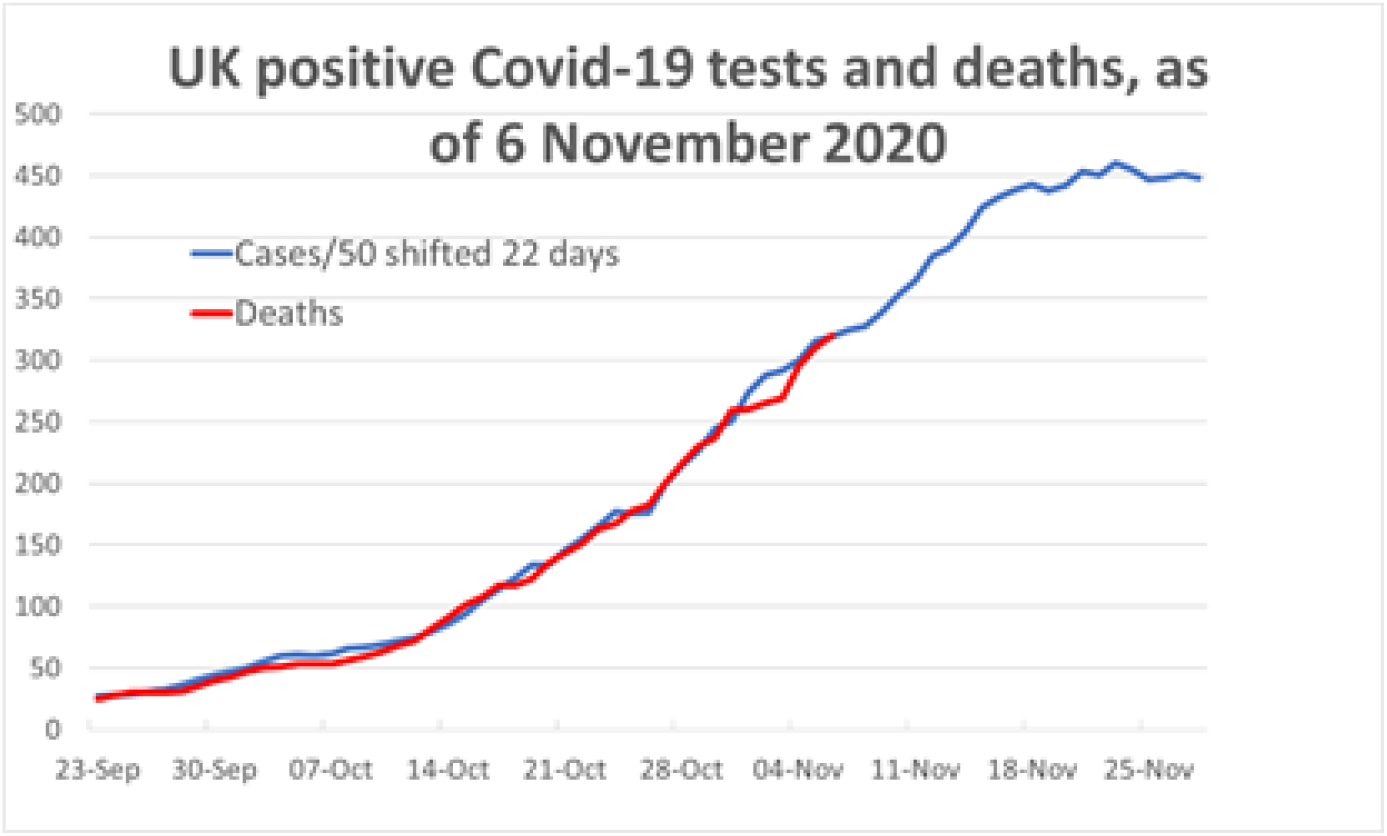
Prediction of deaths from cases during early phase of second wave.

**Figure 2.**
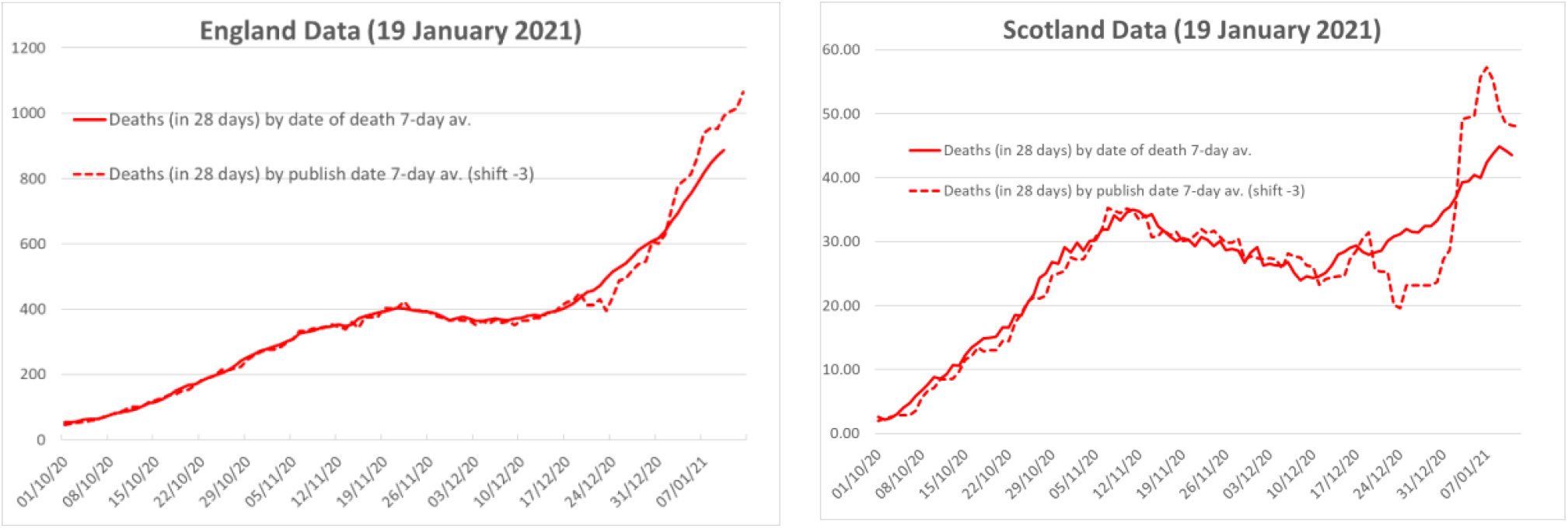
Reported deaths in England and Scotland, 7 day averages. Total deaths - the area under the graph – must eventually be equal independent of whether actual date of death or published date is used. The published deaths are shifted back 3 days in the comparison, recognising a statistical delay in their reporting, and that headline deaths published each day are actually accumulated the previous day. There was clearly significant under-reporting over the Christmas/ New Year period. The effect is particularly marked in the Scotland data.

The headline figure published daily for positive tests daily can be misleading for the same reason, and we henceforth use only the 7-day average of the number of tests by the date the test was taken, i.e. by ‘specimen date’.

In addition to UK government test data, the Office for National Statistics (ONS) monitors prevalence in a representative fixed cohort. This gives us an alternative measure of the number of cases, free from the uncontrolled and unrepresentative testing in the national data, but limited by a much smaller sample size.

In this work we will study whether a two-parameter “scale and shift” model can reliably predict subsequent deaths from reported positive cases, how this varies across regions and whether the VOC impacts the predictions.

Because we are dealing with a *ratio* of tests to cases, a “scale and shift” relationship is unaffected by the total number of cases, so it should work as well regardless of intervention to reduce spread, and for the more infectious VOC provided tests are equally effective and morbidity is similar to the original strain.

Testing is believed to be equally effective for the VOC. A matched comparator (Public Health England Dec. 2020) study of 1,769 VOC and wild type cases showed fewer VOC hospital admissions 16/26 and more VOC deaths 12/10: neither of these is statistically significant, so there is no current evidence to suggest the VOC is associated with higher mortality [1].

## 3 Methods

We address two issues which arise if one simply takes the total number of positive cases, and shifts them forward by a certain number of days, representing an average of time to death. First there is a range of dates from positive test to death, up to the maximum of 28 days now used in Government reporting. By averaging both the cases and deaths over 7 days, we are implicitly allowing a distribution of days to death around the average - in fact, the triangular distribution (1,2,3,4,5,6,7,6,5,4,3,2,1)/49. This is imperfect, but we do it in the absence of specific information on the actual distribution of days to death. We have explored other distributions. They do not appear to modify the broad conclusions of this report.

Second, following a positive test, the likelihood of death is extremely small for the young, and increases rapidly with age. So the likely number of deaths following any given day of tests depends on the age distribution of those who have tested positive. For England, the data on positive cases by age can be downloaded from a dashboard [2]. The Centre for Disease Control in the USA gives information weekly which enables an estimate of the likelihood of death according to age[3], and CDC has taken data from August to estimate the relative likelihood of Covid-related death according to age [4]. The data for ages between 20 and 90 fit an exponential curve with a 2.59 increase in likelihood of death every 10 years. We weight the number of cases by this age-dependent likelihood of death in the ‘CDC weighted’ data below, and postulate that the deathrate is more strongly correlated with this data than the simple case numbers.

Our aim is to collect in a single chart the data for England covering deaths, positive cases, hospital admissions and virus tests, in the way that deaths and cases were shown on the chart 1. We also investigate the weekly data from the Office for National Statistics (ONS) Infection Survey including 95% credible error estimates [5].

For every series of data, we fix the shift and scale factor to agree with the first peak of National 7-day average deaths, 404, on 21 November. This uniquely defines the parameters, implicitly assuming that any reporting delays are equivalent across the country; it does not require or imply that the peak occurs on the same day in all areas. We do not expect such simple shifting and scaling to capture the immense complexity of the processes that underlie the data, so meaningful conclusions can only be drawn from strong signals.

### 3.1 Patient and public involvement

Patients or the public were not involved in the design, or conduct, or reporting, or dissemination plans of our research. All data used was retrieved from existing, public sources as referenced.

## 4 Results

In describing the results we will refer to positive tests results as “cases” and the ratio with deaths as the “case fatality ratio” (CFR). This is significantly larger than the epidemiologically familiar “infection fatality ratio” because many infections are insufficiently severe to trigger a test, and so never detected. Provided the fraction of untested cases remains constant, this will not cause any change in CFR with time, regardless of total numbers. Exactly the same argument applies to the fraction of false positive tests.

The fit to the November peak gives a case-fatality ratio of 1:55. The cases by specimen date which are age-weighted from CDC data as described above to fit the November peak with the following likelihoods of death:

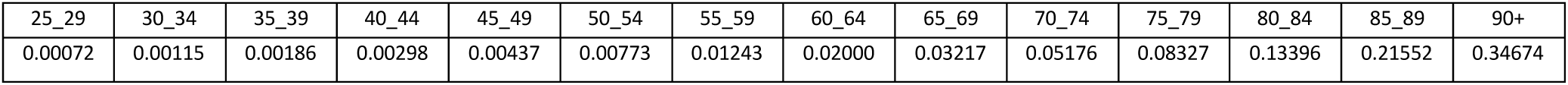

It is immediately clear that, as anticipated, the raw case data (blue line) does not fit the deaths (red line) either before or after the peak on 10 November as well as the cases weighted by age. The early excess when using unweighted cases arises because early in the second wave cases were predominantly in younger age groups.

The data for hospital admissions can also be shifted and scaled to the November peak, and are also shown: we note that the admissions data includes only those who are hospitalized, and this may be affected by limited capacity. Deaths include those who die outside of hospital as well as inside.

In addition to UK government test data, the Office for National Statistics (ONS) monitors prevalence in a representative fixed cohort. This gives us an alternative measure of the number of cases, free from the uncontrolled and unrepresentative testing in the national data, but limited by a much smaller sample size.

Figure 4 shows the results using the ONS infection survey in place of positive tests, including the 95% credible estimates. As with the raw case data, the prediction for the deaths data is poor both before and after the November peak. This also proves that the shift-and-scale procedure does not inevitably give a good fit. However, the age-weighted positive test data for October and November are in remarkable agreement with subsequent deaths.

**Figure 3.**
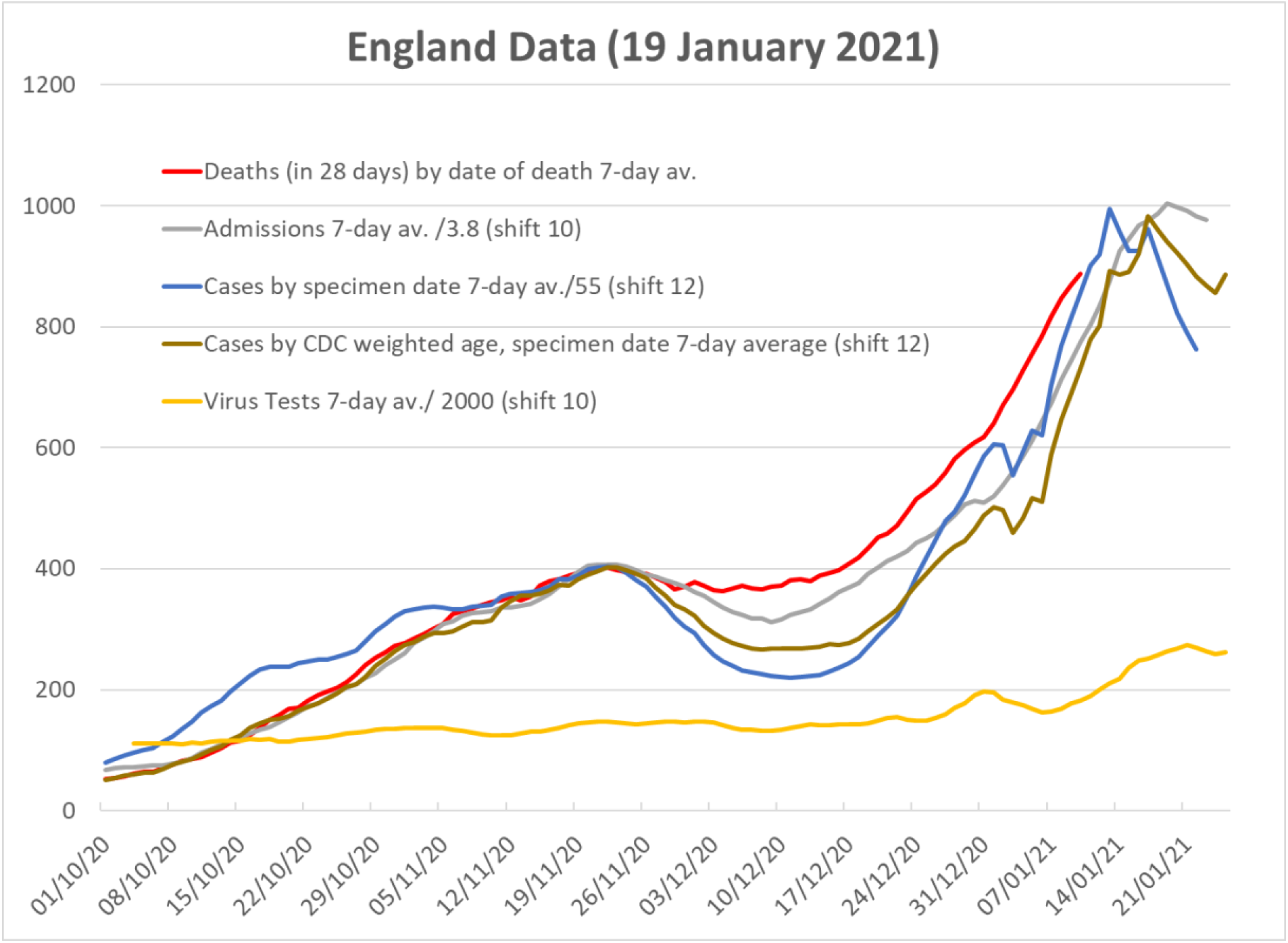
Data for England for deaths, compared with shifted and scaled data for admissions, raw data for (positive test) cases, cases weighted to account for age-morbidity. The shift-and-scale factors are set so that all curves are coincident at the November peak. The yellow line shows the total number of tests.

**Figure 4.**
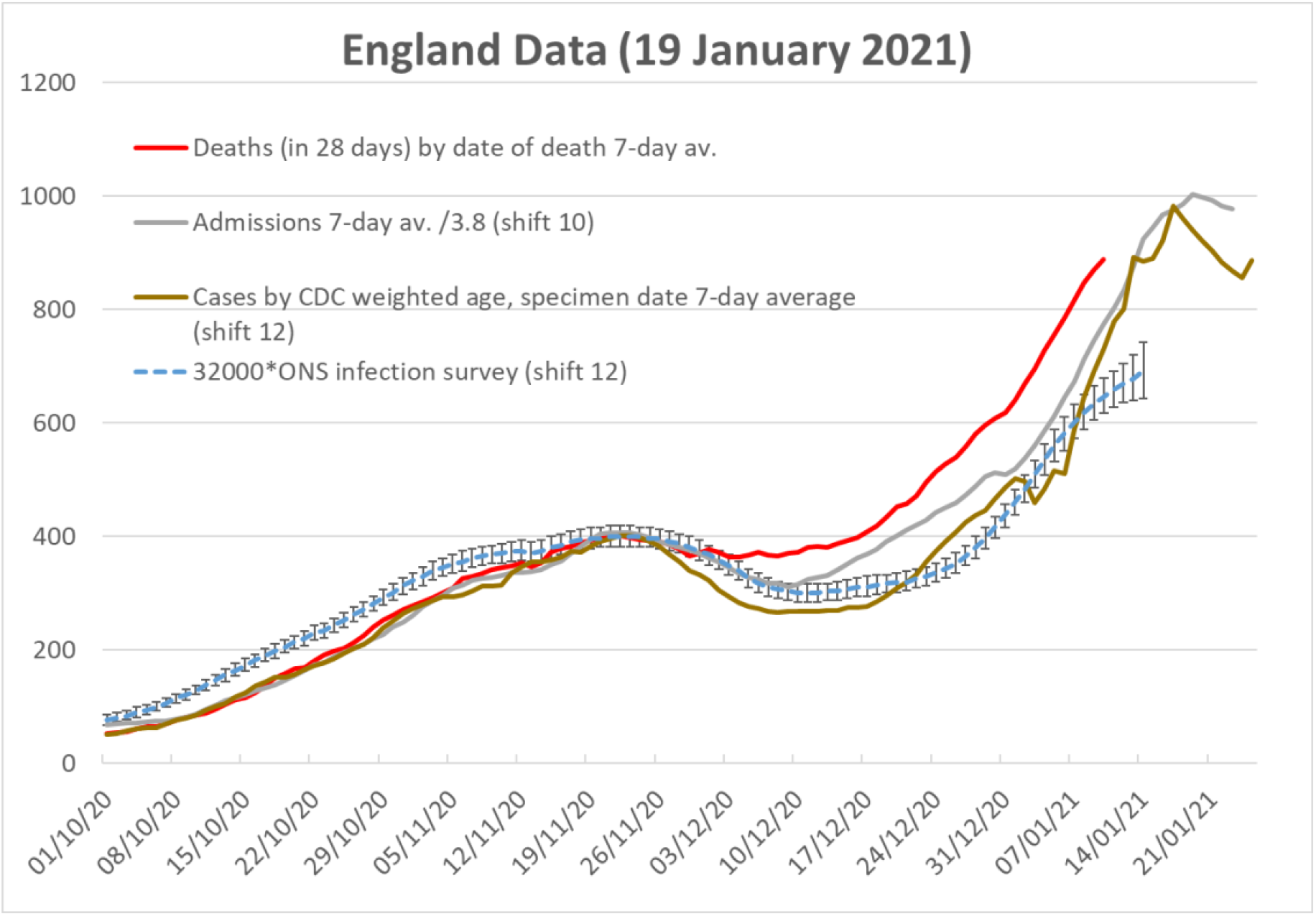
similar to Fig 3, also showing cases predicted from ONS cohort survey, appropriately age-weighted, shifted and scaled to fit the November peak of deaths.

In December the model fails: the dip in cases and admissions has not been reflected in a dip in deaths. This increase in observed over expected deaths demonstrates that something has changed. The continuing steady rise in the number of Virus Tests (yellow line in Figure 3) through December suggests that this is not an artefact from the undertesting.

Having parameterised the model on UK-wide data, we tested its tentative conclusions in different regions. Two cases are described here in detail.

Figure 5 shows data for Yorkshire and Humberside. It shows that the estimate of deaths from shift-and-scaled cases follows the actual deaths beyond the November peak, until the third week of December when an excess of deaths is observed.

**Figure 5.**
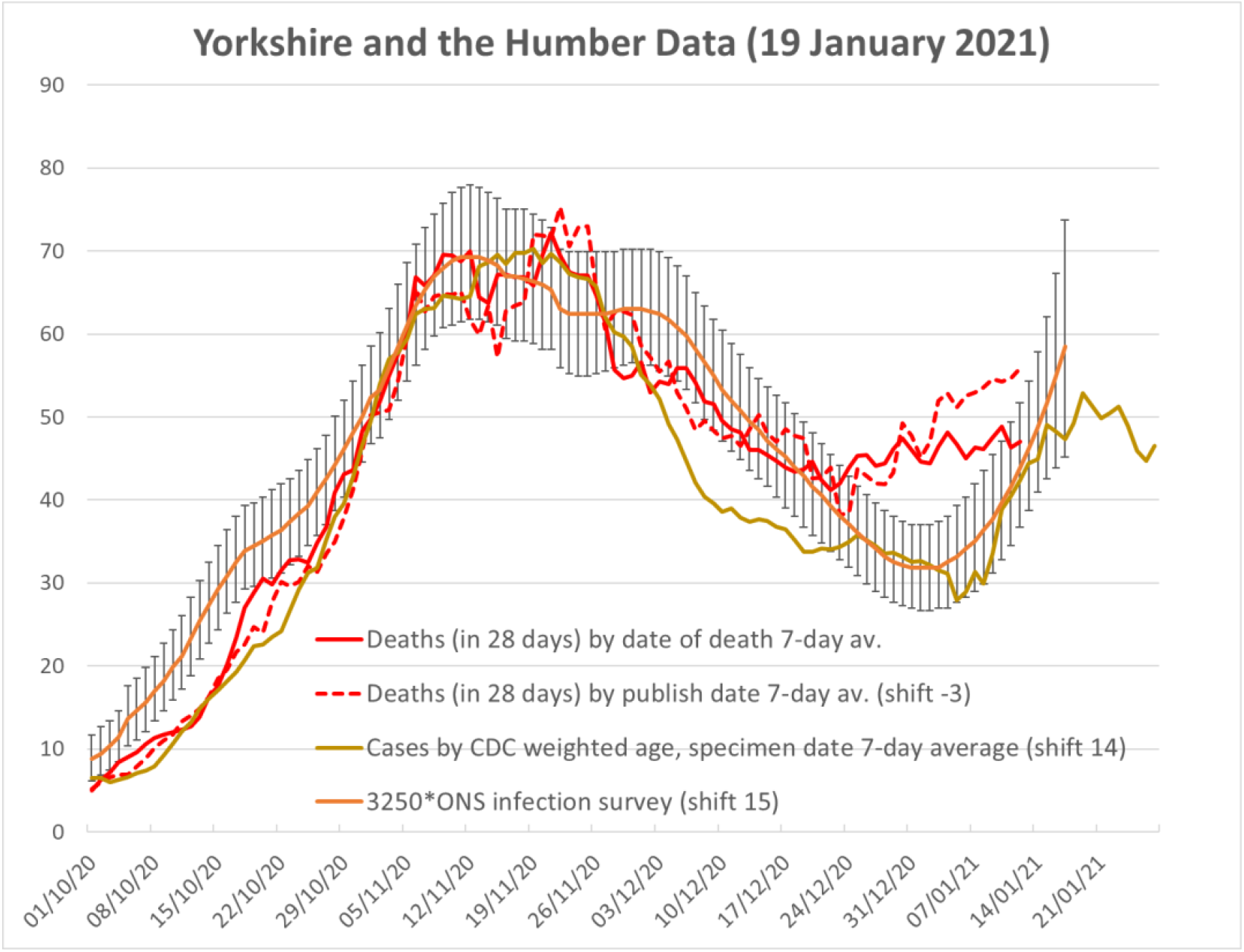
As Figure 4, using data for Yorkshire and Humberside only.

Figure 6 shows data for London. The shape of the curves is quite different from the all- England data. In the chart we have chosen to fix these parameters at the shoulder around 26 November. With this choice, the shifts required are greater than for England as a whole. If this were true, it would imply longer times from specimen date of test to possible admission and death, but other choices of shift cannot be ruled out. Compared with the England scale factor of 32000 for the ONS data, the London scale factor of 3100 is only about 60% of what one would expect from the relative populations of London and England. This demonstrates a consistently lower CFR in London than elsewhere in the UK. i.e. Londoners who test positive are far more likely to survive than those of similar age elsewhere – whether this arises from greater detection of mild cases or better treatment of serious ones is undetermined.

**Figure 6.**
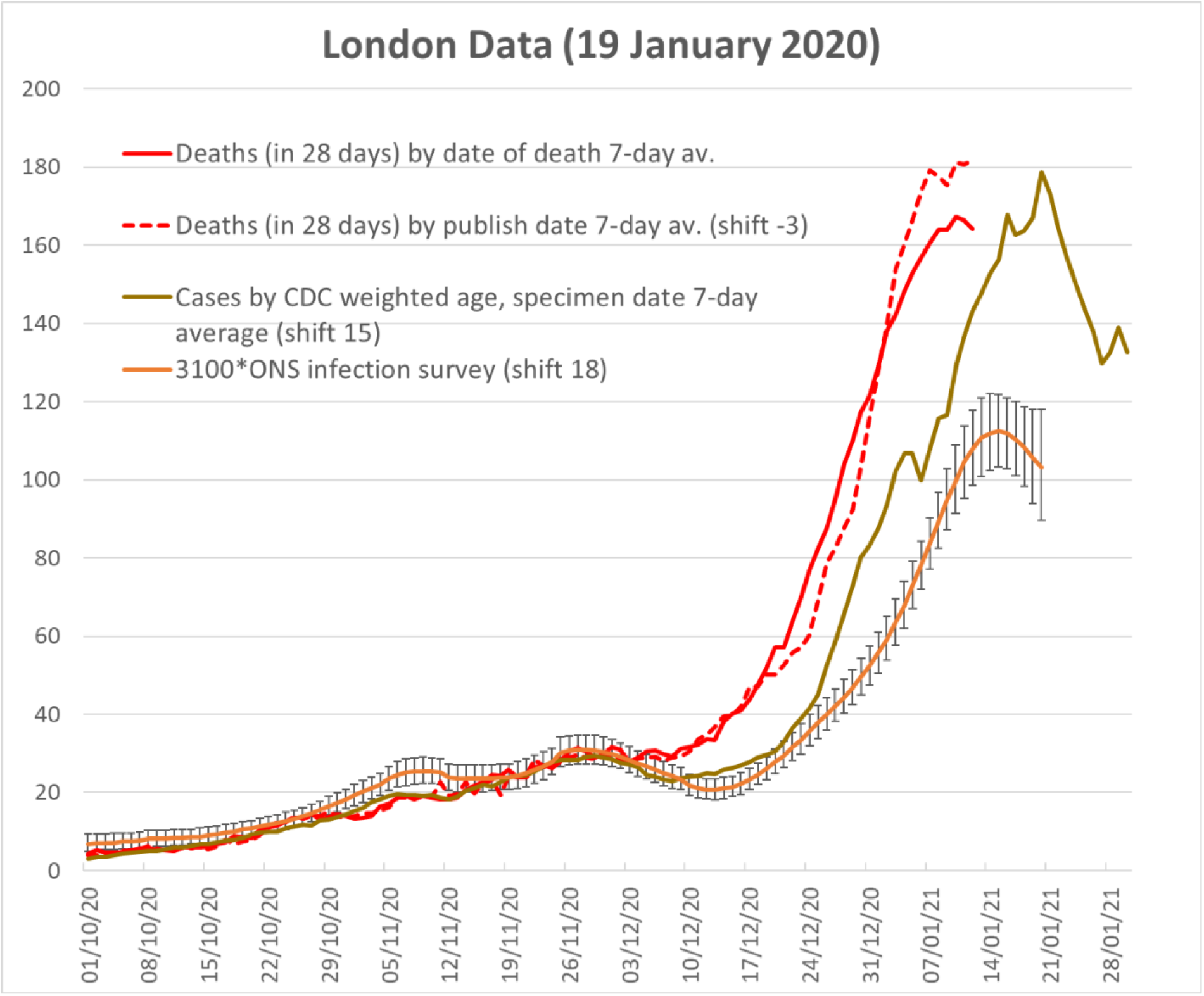
As Figure 4, for London only.

The trend in time for London is similar to the UK, with deaths exceeding the scale-and-shift prediction after late November.

## 5 Discussion

What are the possible explanations of why deaths since the beginning of December have been higher than expected from the number of positive tests?

A higher morbidity for the VOC is one among many possible reasons. Its appearance at a significant level in the South East in December matches the timing of the increase in expected deaths. This increase is delayed in regions such as Yorkshire where the VOC arrived later.

Since 22 November, ONS have been publishing statistical data on the prevalence of ‘new variant compatible’ in positive tests’. The disagreements between test-revealed cases and subsequent deaths in the charts above show qualitatively similarity to the growth in the incidence of the new variant reported by ONS.

In figure 7, we plot the excess of deaths divided by the corresponding prediction from new cases (CDC weighted, by specimen date, 7-day average, shifted by 12, as in Figure 3). As expected from Figure 3, it is roughly 1 up to the beginning of December, and increases thereafter to around 1.3 to 1.4. The blue line uses ‘new variant compatible’ data from ONS [6]

**Figure 7.**
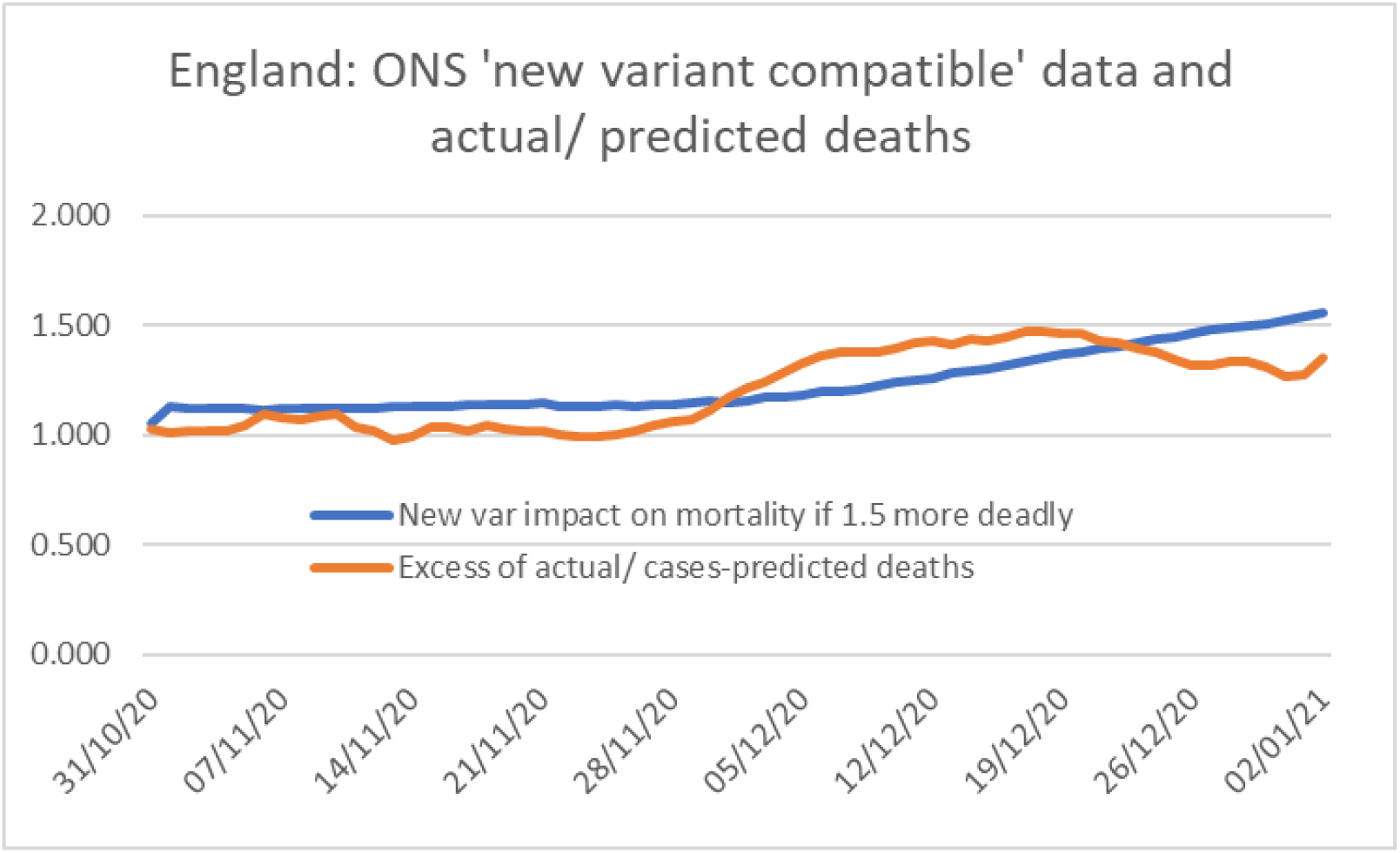

In a release on 20 December Public Health England reported that there was currently no evidence that the new variant ‘VUI – 202012/01’ is more likely to cause severe disease or mortality [7]. Simply for illustrative purposes, the chart shows the effect of assuming the ‘new variant compatible’ has mortality which is 1.5 times greater than ‘other’ – a value which falls within the uncertainty of the PE study. The general increasing trend is similar trend, but the dip in CFR over the Christmas period occurs in spite of increases VOC prevalence. We conclude that the new variant cannot explains the discrepancy between test-revealed cases and subsequent deaths in this period.

Another possible explanation is that not enough tests were being done in December, or the focus of the tests changed to groups less likely to be infected. The first idea is not supported by the yellow line showing the 7-day average of the number of tests (divided by 2000 to fit comfortably on the chart). To explain the CFR increase would require the case detection to get worse, even while the number of tests is increased. It is notable that the ONS cohort data in figure 3 is significantly higher than the age weighted positive test data, which could point to undertesting in the latter.

A third possibility is that the reported deaths within “28 days of a positive test” include a significant number of “normal” winter deaths, and should not be attributed to coronavirus at all. Analysing this is confounded by the abnormally low levels of influenza – a major contributor to “normal” winter excess deaths.

Finally, we mention that the time shifts we employed seem sensible. For example, the extensive analysis by the Imperial group [8] gives average times from hospital admission to death in various circumstances: roughly 10 days average if in a general ward, 12 if in an MV bed following 5 in a general ward, and more for the small number who die after ‘stepping down’ from an MV bed. The rough analysis using the charts on p. 7 of this report suggest an average time from admission to death for those admitted to hospital may be around 12 days. We have not been able to find relevant data on the progression of disease outside of hospital, e.g. in Care Homes.

## 6 Conclusion

We have shown that the relationship between positive tests and subsequent deaths in the UK change significantly around early December, such that the shift-and-scale relationship between test-revealed cases and subsequent deaths failed. The model is fully determined by data, so this observation means there was a marked increase in the case-fatality ratio, where cases (positive tests) and fatalities are as defined by the UK government.

The correlation of increased case-fatality rate with the VOC suggests possible causation. We can identify several possibilities. One interpretation would be that the new variant of the coronavirus produces a more severe infection, as well as being more transmissible, however there is currently no clinical evidence for that and the dip in the CFR in late December does not support this hypothesis; alternately, the test may be less sensitive to the VOC, or the VOC infection may have taken off in undertested parts of the population.

## Data Availability

All data is publicly available via references provided in the paper

## Contributors

DJW designed the analysis for the project, collected the data, performed the analysis and created the figures. GJA suggested comparing regions with different VOC B.1.1.7 prevalence. DJW and GJA wrote the paper together. We thank Mike Cates and David Spiegelhalter for encouraging comments.

## Patients and Public statement

Patients or the public were not involved in the design, or conduct, or reporting, or dissemination plans of our research.

## Ethical approval

No ethical approval was required for this research.

## Data sharing

All data used in this project is available in public domain as referenced

## Transparency

The lead author affirms that the manuscript is an honest, accurate, and transparent account of the study being reported; that no important aspects of the study have been omitted; and that any discrepancies from the study as planned have been explained.

## Dissemination to participants

Since this research uses public demographic data for the whole of the UK, there are no plans for dissemination of this research to specific participants, beyond publishing it.

